# SARS-CoV-2 Seroprevalence Survey Among District Residents Presenting for Serologic Testing at Three Community-Based Test Sites — Washington, DC, July–August, 2020

**DOI:** 10.1101/2021.02.15.21251764

**Authors:** Adrienne Sherman, Jacqueline Reuben, Naomi David, Delores P. Quasie-Woode, Jayleen K. L. Gunn, Carrie F. Nielsen, Patricia Lloyd, Abraham Yohannes, Mary Puckett, Jo Anna Powell, Sarah Leonard, Preetha Iyengar, Fern Johnson-Clarke, Anthony Tran, Matthew McCarroll, Pushker Raj, John Davies-Cole, Jenifer Smith, James A. Ellison, LaQuandra Nesbitt

## Abstract

**Background:** The District of Columbia (DC), a major metropolitan area, continues to see community transmission of SARS-CoV-2. While serologic testing does not indicate current SARS-CoV-2 infection, it can indicate prior infection and help inform local policy and health guidance. The DC Department of Health (DC Health) conducted a community-based survey to estimate DC’s SARS-CoV-2 seroprevalence and identify seropositivity-associated factors.

**Methods:** A mixed-methods cross-sectional serology survey was conducted among a convenience sample of DC residents during July 27**–**August 21, 2020. Free serology testing was offered at three public test sites. Participants completed an electronic questionnaire on household and demographic characteristics, COVID-like illness (CLI) since January 1, 2020, comorbidities, and SARS-CoV-2 exposures. Univariate and bivariate analyses were conducted to describe the sample population and assess factors associated with seropositivity.

**Results:** Among a sample of 671 participants, 51 individuals were seropositive, yielding an estimated seroprevalence of 7.6%. More than half (56.9%) of the seropositive participants reported no prior CLI; nearly half (47.1%) had no prior SARS-CoV-2 testing. Race/ethnicity, prior SARS-CoV-2 testing, prior CLI, employment status, and contact with confirmed COVID-19 cases were associated with seropositivity (P<0.05). Among those reporting prior CLI, loss of taste or smell, duration of CLI, fewer days between CLI and serology test, or prior viral test were associated with seropositivity (P≤0.006).

**Conclusions:** These findings indicate many seropositive individuals reported no symptoms consistent with CLI since January or any prior SARS-CoV-2 testing. This underscores the potential for cases to go undetected in the community and suggests wider-spread transmission than previously reported in DC.

**What is already known on this subject?:** Traditional case-based detection and syndromic surveillance efforts might not identify mildly symptomatic or asymptomatic SARS-CoV-2 infections. This is particularly true among people in the general population who do not have increased risk of severe illness or might not be tested otherwise. Consequently, the true population prevalence of prior SARS-CoV-2 infections might not be known.

**What this study adds?:** A community-based seroprevalence survey conducted in Washington, DC, during July 27**–**August 21, 2020 estimated that 7.6% of the convenience sample had antibodies to SARS-CoV-2, indicating prior infection. At the time of this survey, most of the participants reported that they had not been previously infected with or tested for SARS-CoV-2. These findings highlight both the value of serologic surveillance in complementing other surveillance methods, and the importance of continued prevention and mitigation measures, such as maintaining physical distances of at least 6 feet, avoiding crowds and poorly ventilated spaces, practicing frequent hand hygiene, and wearing face masks properly and consistently around people who do not live with you.

## INTRODUCTION

In December 2019, a novel respiratory virus, Severe Acute Respiratory Syndrome Coronavirus 2 (SARS-CoV-2) emerged in Wuhan, China and has caused widespread illness and death throughout the world(1). Within a year of its emergence, SARS-CoV-2 has caused more than 2.4 million deaths worldwide and more than 485,000 deaths in the United States(2). SARS-CoV-2, the virus that causes coronavirus disease 2019 (COVID-19), was first identified in the District of Columbia (DC) on March 7, 2020 in a traveler returning from a conference in Louisville, Kentucky(3). As of February 14, 2021, a total of 39,001 laboratory**-**confirmed cases and 980 associated deaths among DC residents have been reported to the District of Columbia Department of Health (DC Health)(4).

Serology tests, or antibody tests, detect the presence of antibodies in blood. Antibodies are made by a person’s immune response to infections such as SARS-CoV-2. Immunoglobulin (Ig) antibody tests, such as IgG and IgA tests, are particularly useful in detecting prior infections. While nucleic acid amplification tests (NAATs) detect infection at a specific point in time and, when aggregated, indicate the community’s current burden of disease, serologic testing can provide a more complete picture by identifying previously undetected infections. Several studies have demonstrated serology tests’ utility in confirming prior SARS-CoV-2 infection among people with suspected cases that did not test positive by reverse transcription polymerase chain reaction (RT-PCR) during their illness(5). In such cases, it is possible that the viral load at the time of sample collection was insufficient to reach the detection threshold, yielding false-negative results. Alternatively, incorrect collection, poor storage conditions, or incorrect sample processing could cause inaccurate RT-PCR test results. Additional studies have found the combined use of serologic assays with NAATs provides a more complete surveillance picture(5-8).

Jurisdictions across the U.S. have conducted SARS-CoV-2 seroprevalence assessments, but the DC community’s seroprevalence had not yet been evaluated(8-17). Many existing seroprevalence surveys focus on high-risk groups, such as healthcare workers or long-term care residents, while others lack detailed exposure or clinical information typically collected through interviews or questionnaires(8-17). Data on exposure and clinical presentation are crucial to learn about community transmission dynamics among DC’s general population who might not be considered at high risk of infection or severe illness. There also tends to be great variation between studies in the classes of antibodies assessed (i.e., IgG, IgM, IgA). Assay selection is an important study design consideration as these specific immune responses differ in their development timing, longevity, and clinical implications(5, 6, 16-19).

To better understand the extent of the DC community’s experience with and transmission of SARS-CoV-2, DC Health and the DC Department of Forensic Sciences Public Health Laboratory (DFS-PHL) collaborated with the CDC to conduct a serologic survey during July 27**–** August 21, 2020. This assessment aimed to estimate the proportion of DC residents with evidence of prior SARS-CoV-2 infection, and to identify exposures, comorbidities and symptoms associated with seropositivity.

## METHODS

### Study Design and Participants

A mixed-methods approach to a cross-sectional study design was used to assess the seroprevalence of SARS-CoV-2 among DC residents (see supplementary material for additional details). Initially, a two-stage cluster sampling design was implemented to obtain a representative sample of 560 randomly selected households (Figure 1). Randomly selected households were contacted by mail. However, due to low response, questionnaire and serology test data were concurrently collected from a convenience sample of consenting people who came to any of the testing sites and were not selected through random household sampling. DC residents were made aware of this free testing opportunity and asked to participate through various advertising and outreach strategies (see supplementary material). Data collected from participants recruited from randomly selected households were grouped together with data collected from the convenience sample, and ultimately treated as part of the convenience sample in analyses. This activity was reviewed by DC Health IRBPH and CDC, and was conducted consistent with applicable federal law and CDC policy^§^.

A household member was defined as anyone who spent two or more nights per week in the home. The minimum age for serology testing was six years. Actively ill or symptomatic people were excluded. Households invited to participate as part of the random household sample were offered transportation and one $25 gift card per household. Only participants who completed the questionnaire and the blood draw were enrolled; participants with incomplete questionnaires or invalid serologic results were excluded from analyses. All participants were enrolled during July 27–August 21, 2020 (Supplemental Figure 1).

### Survey Methodology

A standardized questionnaire was developed using Survey123 for ArcGIS and administered to participants on iPads. The questionnaire collected information on household and demographic characteristics, medical history, illnesses and associated symptoms since January 1, 2020, potential SARS-CoV-2 exposures, and prior SARS-CoV-2 testing. Reported symptoms were used to determine whether participants reported a COVID-like illness (CLI) using the Council of State and Territorial Epidemiologists (CSTE) COVID-19 case definition and clinical criteria(20). Participants completed the questionnaires on-site prior to entering the trailer for serology testing.

### Serologic Testing

DC Health established three serology test sites at fixed locations throughout the city (Supplemental Figure 2). Clinicians collected serum specimens from consenting participants using standard venipuncture technique. Serum samples were centrifuged on-site within an hour of collection and refrigerated at 2°C–8°C following centrifugation. Specimens were transported by courier to the DC DFS-PHL for testing, which was completed within 48 hours of specimen receipt. An IgG assay was selected because IgG antibodies are a more reliable indicator of past infections in both symptomatic and asymptomatic patients and are detectable for longer periods of time relative to other classes of antibodies. Serologic testing was conducted using the DiaSorin LIAISON® XL assay (DiaSorin Inc., Stillwater, MN), a qualitative chemiluminescent assay for determination of IgG antibodies to the spike protein (anti-S1 and anti-S2) of SARS-CoV-2. This assay was determined to be highly sensitive (97%) at >14 days post-symptom onset (PSO), and was used under the United States Food and Drug Administration (FDA) Emergency Use Authorization (EUA)(21).

Only negative and positive results for SARS-CoV-2 serology tests were reported to DC Health in accordance with DC Municipal Regulations Chapter 22B 201.1(ff) and 201.1(gg). Participants received their serology results by mail.

### Statistical Analysis

Survey data were matched and merged with participants’ laboratory data. A descriptive analysis was carried out to compare characteristics of respondents to the random household sample with that of the convenience sample. Seroprevalence estimates of SARS-CoV-2 IgG antibodies were calculated as unweighted proportions; corresponding 95% confidence intervals were estimated using the exact binomial test. Descriptive statistics were calculated as crude and relative frequencies for categorical variables; medians and interquartile ranges were calculated for continuous variables. Differences in demographic, clinical and other descriptive characteristics between participants with (seropositive) and without (seronegative) SARS-CoV-2 IgG antibodies were assessed using Wilcoxon rank-sum tests for continuous variables, and Fisher’s exact tests or Pearson’s chi-squared tests for categorical variables. Statistical analyses were two-sided, with significance defined at the 0.05 level. All analyses were conducted using SAS (version 9.4; SAS Institute).

## RESULTS

The final analytic sample consisted of 508 households and 671 participants, averaging 1.3 participants per household (Figure 1). Among 879 randomly selected households initially approached through mailed invitations, 100 (11%) households were enrolled. The analytic sample included 99 randomly selected households, attaining 18% of the targeted 560 randomly selected households. Participants from randomly selected households accounted for 156 of 671 participants.

Descriptive analyses of participants selected through random household sampling compared with participants obtained through convenience sampling identified significant differences in the distributions of age (*p*=0.016), prior testing for SARS-CoV-2 (*p*=0.002), CLI since January 1^st^ (*p*=0.001), self-reported employment status and location (*p*=0.005), contact with confirmed COVID-19 case(s) (*p*=0.015), contact with any person(s) with respiratory symptoms, but unconfirmed COVID-19 (*p*=0.030), and international travel since January 1^st^ (*p*=0.015) (Supplemental Table 1). While not significant, a higher percentage of non-Hispanic Black participants was observed in the random household sample compared with the convenience sample (25% vs. 20% respectively). The seroprevalence point estimate among the random household sample was lower than the seroprevalence among the convenience sample, although the 95% CI overlapped (3.2%, [95% CI 1.1, 7.3] vs. 8.9% [95% CI 6.6, 11.7]) (Supplemental Table 2).

Table 1 presents descriptive characteristics for our sample and the DC population. The majority (60%) of participants identified as Non-Hispanic White, and almost half (48%) were aged 30–49 years. Half (50%) of participants reported working from home some or most days, while 22% of participants reported working outside the home some or most days. Forty percent of participants indicated any prior testing for SARS-CoV-2, and nearly 1/3 (30%) reported symptoms consistent with a CLI since January 1, 2020.

**TABLE 1:** Characteristics of District residents tested for SARS-CoV-2 IgG antibodies by a community-based seroprevalence survey — Washington, DC, July 27–August 21, 2020.

| Characteristic | **Overall Sample (N = 671)** |  | **DC Population (N = 705,749)<sup>†</sup>** |  |
| --- | --- | --- | --- | --- |
|  | No. | Percent* | No. | Percent* |
| ***Gender*** |  |  |  |  |
| Female | 359 | 53.5 | 371,224 | 52.6<sup>†</sup> |
| Male | 312 | 46.5 | 334,525 | 47.4<sup>†</sup> |
| ***Race/Ethnicity*** |  |  |  |  |
| Hispanic ethnicity | 54 | 8.1 | 79,750 | 11.3<sup>†</sup> |
| Black, non-Hispanic | 141 | 21.0 | 311,235 | 44.1<sup>†</sup> |
| White, non-Hispanic | 403 | 60.1 | 263,244 | 37.3<sup>†</sup> |
| Other, non-Hispanic | 52 | 7.8 | 50,814 | 7.2<sup>†</sup> |
| Other, unknown | 21 | 3.1 | -- | -- |
| ***Age Group (yrs)\*\**** |  |  |  |  |
| 6–17 | 29 | 4.3 | 74,838<sup>§</sup> | 10.6 |
| 18–29 | 125 | 18.6 | 155,080<sup>§</sup> | 22.0 |
| 30–49 | 321 | 47.8 | 229,458<sup>§</sup> | 32.5 |
| 50–64 | 132 | 19.7 | 105,700<sup>§</sup> | 15.0 |
| ≥65 | 64 | 9.5 | 87,343<sup>§</sup> | 12.4 |
| ***Prior Testing for SARS-CoV-2*** |  |  |  |  |
| Yes | 265 | 39.5 | 124,027<sup>¶</sup> | 17.6 |
| ***COVID-Like Illness<sup>§§</sup> since January 1, 2020*** |  |  |  |  |
| Yes | 204 | 30.4 | -- | -- |
| ***Medical History*** |  |  |  |  |
| Any chronic condition<sup>†</sup> | 180 | 26.8 | -- | -- |
| Chronic lung disease | 101 | 15.1 | -- | -- |
| Cardiovascular disease | 81 | 12.1 | -- | -- |
| Chronic kidney disease | 8 | 1.2 | -- | -- |
| Liver Disease | 8 | 1.2 | -- | -- |
| Diabetes mellitus | 16 | 2.4 | -- | -- |
| Autoimmune, Rheumatologic, or Immunocompromising condition | 23 | 3.4 | -- | -- |
| Seasonal allergies | 368 | 54.8 | -- | -- |
| Recent/current pregnancy | 6 | 0.9 | -- | -- |
| ***Current Employment Status*** |  |  |  |  |
| Unemployed/furloughed | 95 | 14.2 | -- | -- |
| Employed outside the home | 146 | 21.8 | -- | -- |
| Employed and teleworking | 334 | 49.8 | -- | -- |
| Retired | 55 | 8.2 | -- | -- |
| Student or <18 years of age | 41 | 6.1 | -- | -- |
| ***Exposures since January 1, 2020*** |  |  |  |  |
| No known exposures | 415 | 61.9 | -- | -- |
| Contact with ≥1 person with confirmed COVID-19 | 80 | 11.9 | -- | -- |
| Contact with ≥1 person with respiratory symptoms (not confirmed COVID-19) | 106 | 15.8 | -- | -- |
| International travel (outside the United States) | 116 | 17.3 | -- | -- |
| **Number of Participating Household Members** |  |  |  |  |
| 1 | 381 | 56.8 | -- | -- |
| 2 | 208 | 31.0 | -- | -- |
| 3 | 39 | 5.8 | -- | -- |
| 4 | 28 | 4.2 | -- | -- |
| 5 | 15 | 2.2 | -- | -- |

**Abbreviations:** IgG = Immunoglobulin G; DC = District of Columbia; No. = Number; CI = Confidence Interval;
\*Percentages for mutually exclusive variables might not total 100.0 due to rounding.
<sup>1</sup>Data Source: U.S. Census Bureau; 2019 American Community Survey 1-Year Estimates, Table CP05; generated using data.census.gov; <<https://data.census.gov/cedsci/>>; (6 February 2021).
<sup>5</sup>Data Source: U.S. Census Bureau; Annual Estimates of the Resident Population by Single Year of Age and Sex for District of Columbia: April 1, 2010 to July 1, 2019 (SC-EST2019-SYASEX-11); <<https://www.census.gov/data/datasets/time-series/demo/popest/2010s-state-detail.html>>; (6 February 2021).
<sup>†</sup>Data Source: Government of the District of Columbia; COVID-19 Surveillance Data; <<https://coronavirus.dc.gov/data>> (6 February 2021).
\*\*Minimum age for serologic testing was 6 years; children aged 0–5 years accounted for the remaining 7.5% (n=53,330) of the DC population
<sup>§§</sup>An illness was categorized as compatible with COVID-19 if reported symptoms met the Council of State and Territorial Epidemiologists (CSTE) clinical criteria in the case definition, including (1) cough, difficulty breathing or shortness of breath, new loss of taste, or new loss of smell, or (2) two or more other symptoms (fever [measured or subjective], chills, rigors, myalgia, headache, sore throat, nausea or vomiting, diarrhea, fatigue, congestion or runny nose). https://cdn.ymaws.com/www.cste.org/resource/resmgr/ps/positionstatement2020/Interim-20-ID-02_COVID-19.pdf
<sup>††</sup>Some participants reported multiple chronic conditions; chronic conditions included chronic lung diseases, cardiovascular diseases, chronic kidney diseases, liver diseases, diabetes mellitus, and autoimmune, rheumatologic, or immunocompromising conditions.

The observed seroprevalence of SARS-CoV-2 among 671 participants was 7.6% (95% CI 5.7, 9.9) (Table 2). Positive serology results were observed among 44 (9%) of the 508 distinct households; four households had more than one household member that was found to be seropositive. Seroprevalence estimates were highest among Hispanics (16.7%) and adults aged 18–29 years (11.2%), followed by children aged 6–17 years (10.3%). Differences in seroprevalence estimates for those working outside of the home compared with those employed and teleworking were observed, with a higher seroprevalence among those working outside the home (13.0% [95% CI 8.0, 19.6] vs. 5.1% [95% CI 3.0, 8.0]).

**TABLE 2:**
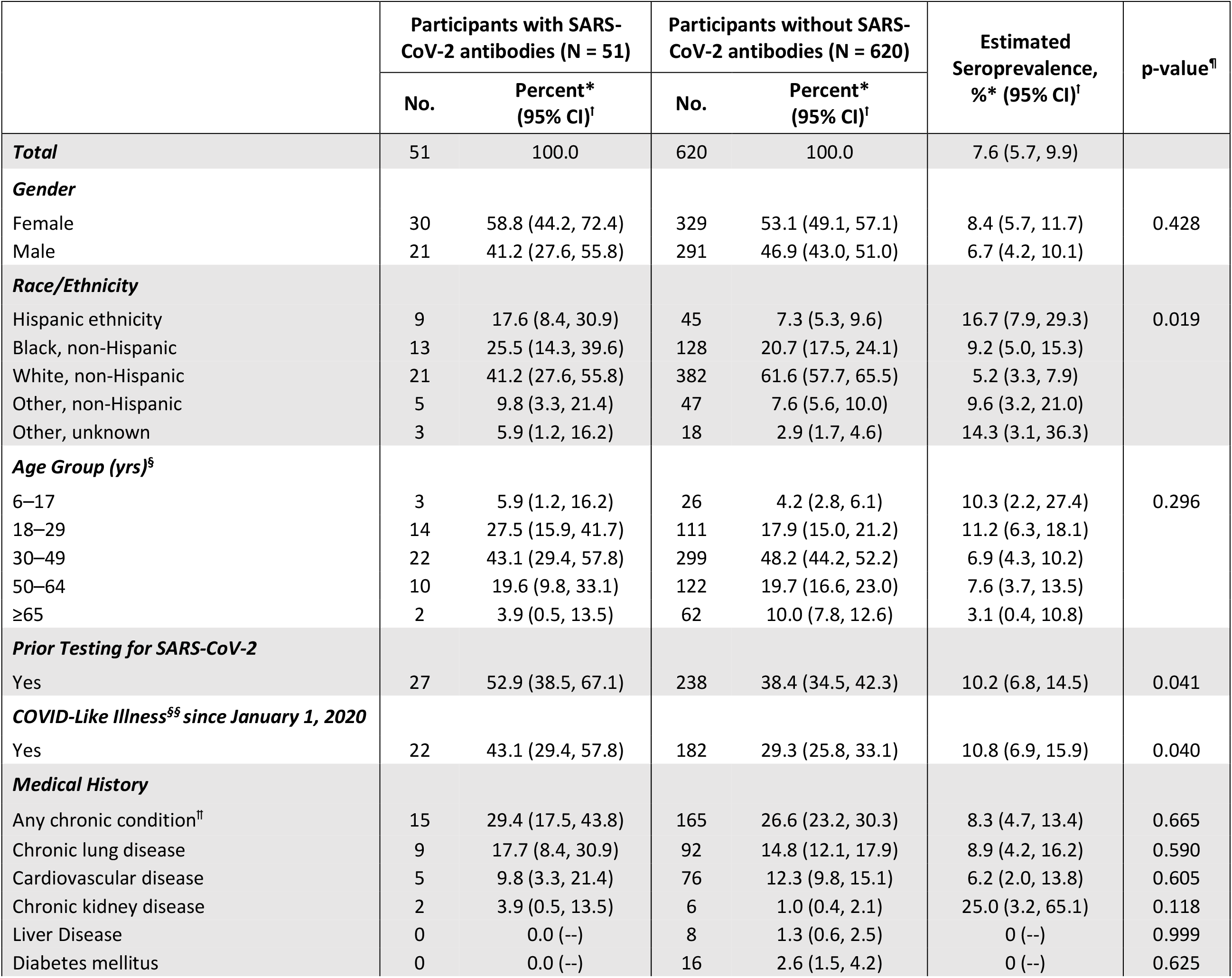
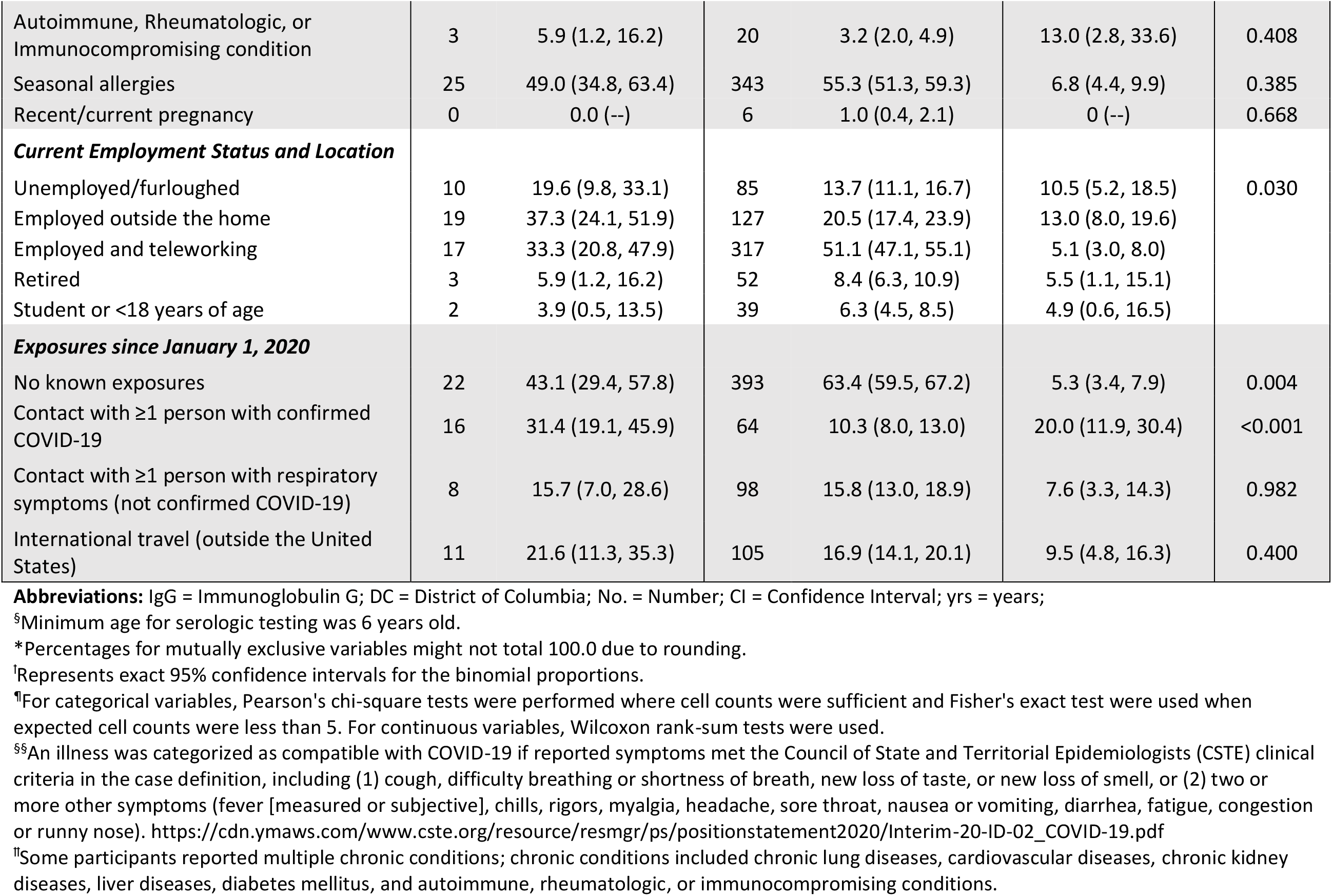
Characteristics of participants with and without SARS-CoV-2 IgG antibodies, estimated seroprevalence, and factors associated with seropositivity — Washington, DC, July 27–August 21, 2020.

|  | Participants with SARS-CoV-2 antibodies (N = 51) |  | Participants without SARS-CoV-2 antibodies (N = 620) |  | Estimated Seroprevalence, %* (95% CI)<sup>†</sup> | p-value<sup>¶</sup> |
| --- | --- | --- | --- | --- | --- | --- |
|  | No. | Percent* (95% CI)<sup>†</sup> | No. | Percent* (95% CI)<sup>†</sup> |  |  |
| **Total** | 51 | 100.0 | 620 | 100.0 | 7.6 (5.7, 9.9) |  |
| **Gender** |  |  |  |  |  |  |
| Female | 30 | 58.8 (44.2, 72.4) | 329 | 53.1 (49.1, 57.1) | 8.4 (5.7, 11.7) | 0.428 |
| Male | 21 | 41.2 (27.6, 55.8) | 291 | 46.9 (43.0, 51.0) | 6.7 (4.2, 10.1) |  |
| **Race/Ethnicity** |  |  |  |  |  |  |
| Hispanic ethnicity | 9 | 17.6 (8.4, 30.9) | 45 | 7.3 (5.3, 9.6) | 16.7 (7.9, 29.3) | 0.019 |
| Black, non-Hispanic | 13 | 25.5 (14.3, 39.6) | 128 | 20.7 (17.5, 24.1) | 9.2 (5.0, 15.3) |  |
| White, non-Hispanic | 21 | 41.2 (27.6, 55.8) | 382 | 61.6 (57.7, 65.5) | 5.2 (3.3, 7.9) |  |
| Other, non-Hispanic | 5 | 9.8 (3.3, 21.4) | 47 | 7.6 (5.6, 10.0) | 9.6 (3.2, 21.0) |  |
| Other, unknown | 3 | 5.9 (1.2, 16.2) | 18 | 2.9 (1.7, 4.6) | 14.3 (3.1, 36.3) |  |
| **Age Group (yrs)<sup>§</sup>** |  |  |  |  |  |  |
| 6–17 | 3 | 5.9 (1.2, 16.2) | 26 | 4.2 (2.8, 6.1) | 10.3 (2.2, 27.4) | 0.296 |
| 18–29 | 14 | 27.5 (15.9, 41.7) | 111 | 17.9 (15.0, 21.2) | 11.2 (6.3, 18.1) |  |
| 30–49 | 22 | 43.1 (29.4, 57.8) | 299 | 48.2 (44.2, 52.2) | 6.9 (4.3, 10.2) |  |
| 50–64 | 10 | 19.6 (9.8, 33.1) | 122 | 19.7 (16.6, 23.0) | 7.6 (3.7, 13.5) |  |
| ≥65 | 2 | 3.9 (0.5, 13.5) | 62 | 10.0 (7.8, 12.6) | 3.1 (0.4, 10.8) |  |
| **Prior Testing for SARS-CoV-2** |  |  |  |  |  |  |
| Yes | 27 | 52.9 (38.5, 67.1) | 238 | 38.4 (34.5, 42.3) | 10.2 (6.8, 14.5) | 0.041 |
| **COVID-Like Illness<sup>§§</sup> since January 1, 2020** |  |  |  |  |  |  |
| Yes | 22 | 43.1 (29.4, 57.8) | 182 | 29.3 (25.8, 33.1) | 10.8 (6.9, 15.9) | 0.040 |
| **Medical History** |  |  |  |  |  |  |
| Any chronic condition<sup>¶¶</sup> | 15 | 29.4 (17.5, 43.8) | 165 | 26.6 (23.2, 30.3) | 8.3 (4.7, 13.4) | 0.665 |
| Chronic lung disease | 9 | 17.7 (8.4, 30.9) | 92 | 14.8 (12.1, 17.9) | 8.9 (4.2, 16.2) | 0.590 |
| Cardiovascular disease | 5 | 9.8 (3.3, 21.4) | 76 | 12.3 (9.8, 15.1) | 6.2 (2.0, 13.8) | 0.605 |
| Chronic kidney disease | 2 | 3.9 (0.5, 13.5) | 6 | 1.0 (0.4, 2.1) | 25.0 (3.2, 65.1) | 0.118 |
| Liver Disease | 0 | 0.0 (–) | 8 | 1.3 (0.6, 2.5) | 0 (–) | 0.999 |
| Diabetes mellitus | 0 | 0.0 (–) | 16 | 2.6 (1.5, 4.2) | 0 (–) | 0.625 |
| Autoimmune, Rheumatologic, or Immunocompromising condition | 3 | 5.9 (1.2, 16.2) | 20 | 3.2 (2.0, 4.9) | 13.0 (2.8, 33.6) | 0.408 |
| Seasonal allergies | 25 | 49.0 (34.8, 63.4) | 343 | 55.3 (51.3, 59.3) | 6.8 (4.4, 9.9) | 0.385 |
| Recent/current pregnancy | 0 | 0.0 (--) | 6 | 1.0 (0.4, 2.1) | 0 (--) | 0.668 |
| **Current Employment Status and Location** |  |  |  |  |  |  |
| Unemployed/furloughed | 10 | 19.6 (9.8, 33.1) | 85 | 13.7 (11.1, 16.7) | 10.5 (5.2, 18.5) | 0.030 |
| Employed outside the home | 19 | 37.3 (24.1, 51.9) | 127 | 20.5 (17.4, 23.9) | 13.0 (8.0, 19.6) |  |
| Employed and teleworking | 17 | 33.3 (20.8, 47.9) | 317 | 51.1 (47.1, 55.1) | 5.1 (3.0, 8.0) |  |
| Retired | 3 | 5.9 (1.2, 16.2) | 52 | 8.4 (6.3, 10.9) | 5.5 (1.1, 15.1) |  |
| Student or <18 years of age | 2 | 3.9 (0.5, 13.5) | 39 | 6.3 (4.5, 8.5) | 4.9 (0.6, 16.5) |  |
| **Exposures since January 1, 2020** |  |  |  |  |  |  |
| No known exposures | 22 | 43.1 (29.4, 57.8) | 393 | 63.4 (59.5, 67.2) | 5.3 (3.4, 7.9) | 0.004 |
| Contact with ≥1 person with confirmed COVID-19 | 16 | 31.4 (19.1, 45.9) | 64 | 10.3 (8.0, 13.0) | 20.0 (11.9, 30.4) | <0.001 |
| Contact with ≥1 person with respiratory symptoms (not confirmed COVID-19) | 8 | 15.7 (7.0, 28.6) | 98 | 15.8 (13.0, 18.9) | 7.6 (3.3, 14.3) | 0.982 |
| International travel (outside the United States) | 11 | 21.6 (11.3, 35.3) | 105 | 16.9 (14.1, 20.1) | 9.5 (4.8, 16.3) | 0.400 |

**Abbreviations:** IgG = Immunoglobulin G; DC = District of Columbia; No. = Number; CI = Confidence Interval; yrs = years;
<sup>§</sup>Minimum age for serologic testing was 6 years old.
\*Percentages for mutually exclusive variables might not total 100.0 due to rounding.
<sup>†</sup>Represents exact 95% confidence intervals for the binomial proportions.
<sup>¶</sup>For categorical variables, Pearson's chi-square tests were performed where cell counts were sufficient and Fisher's exact test were used when expected cell counts were less than 5. For continuous variables, Wilcoxon rank-sum tests were used.
<sup>§§</sup>An illness was categorized as compatible with COVID-19 if reported symptoms met the Council of State and Territorial Epidemiologists (CSTE) clinical criteria in the case definition, including (1) cough, difficulty breathing or shortness of breath, new loss of taste, or new loss of smell, or (2) two or more other symptoms (fever [measured or subjective], chills, rigors, myalgia, headache, sore throat, nausea or vomiting, diarrhea, fatigue, congestion or runny nose). https://cdn.ymaws.com/www.cste.org/resource/resmgr/ps/positionstatement2020/Interim-20-ID-02_COVID-19.pdf
<sup>¶¶</sup>Some participants reported multiple chronic conditions; chronic conditions included chronic lung diseases, cardiovascular diseases, chronic kidney diseases, liver diseases, diabetes mellitus, and autoimmune, rheumatologic, or immunocompromising conditions.

Nearly half (47%) of those with detectable SARS-CoV-2 IgG antibodies indicated they had no prior testing for SARS-CoV-2 (Table 2). Approximately 43% of participants with a positive serology test reported a CLI since January 1, 2020. Non-Hispanic White participants had a lower seroprevalence than all other racial/ethnic groups (Table 2), however, convenience sampling resulted in overrepresentation of non-Hispanic White residents (Table 1). Prior SARS-CoV-2 testing, CLI since January 1st, 2020, contact with any confirmed COVID-19 cases, and employment outside the household were significantly (*p* < 0.05) associated with seropositivity.

Among those with a prior CLI, the median duration from symptom onset to serum specimen collection was significantly longer for seronegative participants compared with seropositive participants (median 163 days vs. 123 days, *p* = 0.001) (Table 3). Seropositivity was significantly associated with loss of taste, loss of smell, illness duration, prior SARS-CoV-2 viral testing, and a prior positive SARS-CoV-2 viral test (*p* < 0.01). Three seronegative participants reporting a prior CLI also reported previously testing positive for SARS-CoV-2 infection.

**TABLE 3:** Characteristics of participants who reported a COVID-like^†^ illness^§^ — Washington, DC, July 27–August 21, 2020.

| Clinical Characteristics | Participants with a COVID-like<sup>1</sup> illness<sup>§</sup> reported (N=204) |  |  |  | p-value<sup>¶</sup> |
| --- | --- | --- | --- | --- | --- |
|  | Participants with SARS-CoV-2 antibodies (N = 22) |  | Participants without SARS-CoV-2 antibodies (N = 182) |  |  |
|  | No. | Percent* (95% CI)<sup>¶</sup> | No. | Percent* (95% CI)<sup>¶</sup> |  |
| ***Reported Symptoms*** |  |  |  |  |  |
| Cough | 12 | 54.6 (32.2, 75.6) | 114 | 62.6 (55.2, 69.7) | 0.461 |
| Chills | 12 | 54.6 (32.2, 75.6) | 75 | 41.2 (34.0, 48.7) | 0.232 |
| Measured fever | 11 | 50.0 (28.2, 71.8) | 58 | 31.9 (25.2, 39.2) | 0.090 |
| *Maximum Fever Temperature, Median (IQR), °F* | 11 | 101.0°F (99.8°F–101.9°F) | 53 | 101.0°F (100.3°F–102.0°F) | 0.271 |
| Subjective fever | 10 | 45.5 (24.4, 67.8) | 72 | 39.6 (32.4, 47.1) | 0.594 |
| Congestion | 8 | 36.4 (17.2, 59.3) | 83 | 45.6 (38.2, 53.1) | 0.410 |
| Sore throat | 9 | 40.9 (20.7, 63.7) | 100 | 55.0 (47.4, 62.3) | 0.213 |
| Chest pain | 3 | 13.6 (2.9, 34.9) | 37 | 20.3 (14.7, 26.9) | 0.579 |
| Muscle pain | 11 | 50.0 (28.2, 71.8) | 54 | 29.7 (23.1, 36.9) | 0.053 |
| Difficulty breathing or short of breath | 6 | 27.3 (10.7, 50.2) | 45 | 24.7 (18.6, 31.7) | 0.794 |
| Abdominal pain | 5 | 22.7 (7.8, 45.4) | 18 | 9.9 (6.0, 15.2) | 0.082 |
| Nausea | 7 | 31.8 (13.9, 54.9) | 28 | 15.4 (10.5, 21.5) | 0.070 |
| Vomiting | 3 | 13.6 (2.9, 34.9) | 17 | 9.3 (5.5, 14.5) | 0.459 |
| Diarrhea | 5 | 22.7 (7.8, 45.4) | 33 | 18.1 (12.8, 24.5) | 0.569 |
| Headache | 14 | 63.6 (40.7, 82.8) | 81 | 44.5 (37.2, 52.0) | 0.089 |
| Fatigue | 15 | 68.2 (45.1, 86.1) | 108 | 59.3 (51.8, 66.6) | 0.423 |
| Loss of taste | 9 | 40.9 (20.7, 63.7) | 21 | 11.5 (7.3, 17.1) | 0.001 |
| Loss of smell | 9 | 40.9 (20.7, 63.7) | 17 | 9.3 (5.5, 14.5) | <0.001 |
| Other Symptom(s) | 5 | 22.7 (7.8, 45.4) | 9 | 5.0 (2.3, 9.2) | <0.010 |
| ***Medical Care Sought for this Illness*** |  |  |  |  |  |
| Any care sought<sup>§§</sup> | 8 | 36.4 (17.2, 59.3) | 71 | 39.0 (31.9, 46.5) | 0.810 |
| **Absences Due to this Illness** |  |  |  |  |  |
| Any absence(s) from work/school | 10 | 45.5 (24.4, 67.8) | 86 | 47.3 (39.8, 54.8) | 0.873 |
| **Visitors During this Illness** |  |  |  |  |  |
| Yes | 2 | 9.1 (1.1, 29.2) | 47 | 25.8 (19.6, 32.8) | 0.112 |
| **SARS-CoV-2 Testing for this Illness** |  |  |  |  |  |
| Viral testing for this illness | 12 | 54.6 (32.2, 75.6) | 32 | 17.6 (12.4, 23.9) | <0.001 |
| Positive viral test result for this illness | 11 | 50.0 (28.2, 71.8) | 3 | 1.7 (0.3, 4.7) | <0.001 |
| **Duration of Illness, Median (IQR), days** | 22 | 14 (7–21) | 175 | 7 (5–14) | 0.006 |
| **Duration from Symptom Onset to Serum Specimen Collection, Median (IQR), days** | 22 | 123 (32–149) | 182 | 163 (135–193) | 0.001 |

**Abbreviations:** DC = District of Columbia; No. = Number; CI = Confidence Interval; IQR = Interquartile range; F = Fahrenheit;
<sup>§</sup>Refers to the most recent illness episode for participants that experienced more than one instance of illness since January 2020.
<sup>†</sup>An illness was categorized as compatible with COVID-19 if reported symptoms met the Council of State and Territorial Epidemiologists (CSTE) clinical criteria in the case definition, including (1) cough, difficulty breathing or shortness of breath, new loss of taste, or new loss of smell, or (2) two or more other symptoms (fever [measured or subjective], chills, rigors, myalgia, headache, sore throat, nausea or vomiting, diarrhea, fatigue, congestion or runny nose). https://cdn.ymaws.com/www.cste.org/resource/resmgr/ps/positionstatement2020/Interim-20-ID-02_COVID-19.pdf
\*Percentages for mutually exclusive variables might not total 100.0 due to rounding.
<sup>††</sup>Represents exact 95% confidence intervals for the binomial proportions.
<sup>¶</sup>For categorical variables, Pearson's chi-square tests were performed where cell counts were sufficient and Fisher's exact test were carried out when expected cell counts were less than 5. For continuous variables, Wilcoxon rank-sum tests were used.
<sup>§§</sup>Includes any telemedicine appointment(s) and any healthcare visit(s) to a doctor, clinic, or emergency room.

## DISCUSSION

Among 671 included participants, SARS-CoV-2 seropositivity was significantly associated with Hispanic ethnicity and non-White race, prior CLI, employment outside of the household, and contact with at least one person with confirmed COVID-19 (*p* < 0.05). The lower seroprevalence observed among people who teleworked compared with those working outside the home suggests an association between telework arrangements and less SARS-CoV-2 exposure or infection. This is consistent with and supportive of findings from a multistate case-control study assessing telework practices in the two weeks preceding SARS-CoV-2 diagnostic testing, which found a significantly lower proportion of people with COVID-19 reporting part- or full-time telework practices compared with controls (35% vs. 53% respectively)(22). Case-patients were also more likely to report regular work or school attendance than control-patients, further highlighting the potential for risk reduction of SARS-CoV-2 infection when employers provide telework options(22).

In our study, almost half (47%) of seropositive participants indicated no prior SARS-CoV-2 testing, and more than half (57%) of seropositive participants reported no symptoms consistent with a CLI since January 1, 2020, which indicate that prior SARS-CoV-2 infections in DC might have gone undetected, likely due to the proportion of asymptomatic or mildly symptomatic cases and the unavailability of testing early in the pandemic. Three seronegative participants reporting a prior CLI reported previously testing positive for SARS-CoV-2 infection, two of whom reported CLI onset 2–4 months prior to serologic testing. This might support other studies’ findings that have demonstrated some people fail to develop anti-SARS-CoV-2 antibodies following infection(5, 18, 23, 24).

Seroprevalence estimates were highest among participants aged 18–29 years, followed by participants aged 6–17 years. While only three participants aged 6–17 years were seropositive, our seroprevalence point estimate for this age group differs from other reports’ findings, as most assessments that included children under 18 years of age have either found no children to be seropositive, or observed children to have a lower seroprevalence relative to most other age groups(17, 25-28). The large proportion of seropositive participants reporting no prior symptoms consistent with CLI might, in part, be explained by the high seroprevalence observed among people aged 6–29 years in our sample, as current literature indicates that younger age correlates with asymptomatic and mild infections(29, 30).

The seroprevalence of SARS-CoV-2 IgG antibodies within a community-based convenience sample of DC residents was estimated to be 7.6% (95% CI 5.7, 9.9). This estimate is consistent with what one would expect for the DC area during this period based on the timing of the first epidemic curve’s peak in DC and the RT-PCR positivity rates in the weeks preceding this serosurvey(4). The seroprevalence observed in DC is higher than estimates observed in other urban areas: 6.9% in NYC during March; 2.4% in Minneapolis-St Paul-St Cloud during May; 1.0% in San Francisco Bay area, 3.2% in Philadelphia, 2.5% in Atlanta, and 4.7% in Los Angeles during April. However, many jurisdictions conducted their seroprevalence assessments earlier in the U.S. epidemic and when many jurisdictions had shelter in place orders to minimize community transmission(14, 25, 26). DC was not under a shelter-in-place order at the time of this survey, rather, DC progressed to phase 2 of reopening on June 22, 2020, which expanded the range of activities and business functions allowed to resume with capacity restrictions in place and permitted reopening of additional businesses and institutions that enable more interactions between community members to take place (i.e., indoor dining at restaurants, outdoor fitness/recreational classes).

Unsurprisingly, the estimated seroprevalence among the random household sample was lower than the seroprevalence among the convenience sample (3.2% vs. 8.9% respectively). Although these seroprevalence estimates did not differ significantly, the higher seroprevalence observed among the convenience sample reflects the anticipated bias of self-selection, as people who think they might have previously been exposed to, or infected with, SARS-CoV-2 were more likely to actively seek out serologic testing. Descriptive analyses between the convenience and random household samples also identified differences in the distributions of age and race/ethnicity, prior CLI, prior SARS-CoV-2 testing, employment status and location, and SARS-CoV-2 exposures. These differences highlight the importance and value of obtaining a representative population-based sample through random sampling to accurately estimate seroprevalence in the community(31).

There are several limitations of this survey. Most notably, the low response rates of mailed invitations led to a reliance on convenience sampling and results are, therefore, not generalizable to the DC population. The use of a convenience sample likely inflated the seroprevalence estimate due to self-selection bias. Test sites’ hours of operation were restricted to weekday business hours, which made participation difficult for those currently employed, particularly in an essential job. The testing sites also remained in fixed locations, which made access less convenient for certain census blocks in DC. Additionally, the current analysis did not account for clustering within a household, as the average number of individuals per enrolled household was 1.3 persons. Inclusion of multiple individuals from the same household was possible, and multiple positive results were observed among 4 of the 44 households with at least one seropositive result. Since IgG antibodies can take 14 days PSO to reach detectable limits, it is possible that persons with an acute infection had not yet produced detectable levels of SARS-CoV-2 antibodies, particularly among people tested within 14 days PSO. Further, as immunological assessments found that not all individuals mount an antibody response following SARS-CoV-2 infection, it is possible that some participants might have had a previous SARS-CoV-2 infection, but did not produce detectable antibodies and, thus, were not identified through serologic testing(5, 18, 23, 24). Lastly, we must emphasize that as of February 7, 2021, the detection of IgG antibodies with a qualitative test might not be at a level that provides protective immunity, and cases of re-infection with SARS-CoV-2, while rare, have already been reported in several countries(24, 32-38).

Despite these limitations, this cross-sectional survey provides additional data for the scientific and public health communities. This is the first assessment of SARS-CoV-2 seroprevalence within the general population of DC. The serology test sites established and operated by DC Health and DFS-PHL were entirely free to the public to improve access to testing among the general public, particularly for uninsured or low-income persons. To incentivize participation among the randomly selected households, all cost and transportation barriers were removed; invited participants were offered transportation vouchers and compensated for their time. Furthermore, the IgG assay implemented here has demonstrated high sensitivity and was used under an EUA from FDA. Despite taking several days PSO to be produced by the body, numerous studies have found SARS-CoV-2 IgG antibody levels decrease at a lower rate over time than other antibody classes (i.e., IgM, IgA), which is advantageous for surveillance purposes(5,18,19).

## Conclusions

This survey collected data on prior illnesses and symptoms consistent with COVID-19, potential SARS-CoV-2 exposures, and medical history that might impact a person’s risk of infection or disease severity. The survey data, when paired with serology results, enables the comparison of stratified seroprevalence estimates, and provides insight on the frequency of SARS-CoV-2 exposures among community members. The observed proportion of seropositive people who reported no symptoms consistent with CLI also provides valuable insight on the transmission dynamics surrounding asymptomatic infections, particularly among younger people observed to have higher SARS-CoV-2 seroprevalence estimates. Further, the higher seroprevalence observed among people who worked outside the home compared with people who worked from home highlights telework policies as a useful non-pharmaceutical intervention to slow the spread of COVID-19 in the community. This information highlights both the value of serologic surveillance in complementing other surveillance methods, and the importance of continued prevention and mitigation measures, even as vaccines and other pharmaceutical interventions become widely available.

## Data Availability

Analyses can be provided upon request. Provision of raw data will not be shared externally outside of DC Health.

## Acknowledgments

The authors wish to acknowledge the following individuals for their vital contributions to this work: Patrick Ashley (DC Health), Health and Medical Branch Director; Travis Cryan (DC HSEMA), COVID-19 Testing and Sampling Group Lead; Dorothy Lowry (DC Health), Antibody Testing and Sampling Team Coordinator; Paul Duray (DC Health), Health and Medical Logistics Lead ; Daniel Burke (DC Health), Health and Medical Branch Assistant Director; Ron Benedict (DC HSEMA), Health and Medical Logistics Support; Statia Thomas (DC Health), Public Health Information Unit Lead; Jennifer Lumpkins (DC Health), COVID-19 Test Result Communications Manager; Todd Jasper, COVID-19 Test Result Communications Manager; Ezra Alltucker (DC HSEMA), Logistical and Operational Support Coordinator; Suparna Das (DC Health) for census block sampling; Ian Quan (DC Health) for geocoding assistance; Timothy Hutchinson for Survey123 for ArcGIS technical support; Nia Deot (DFS-PHL) for providing epidemiology-laboratory coordination and review contributions; Nancy LaVerda (DC Health) for address validation assistance and outreach; Alison Reeves (DC Health) for community outreach efforts and communications support; all members of the DC Health Antibody Sampling and Testing Team, including the Call Center team, who worked tirelessly to schedule and provide testing to thousands of community members.

## Disclaimer

Use of trade names and commercial sources are for identification only and do not imply endorsement by the US Department of Health and Human Services, Centers for Disease Control and Prevention, nor the Government of the District of Columbia. The findings and conclusions in this report are those of the authors and do not necessarily represent the views of their institutions.

## Footnotes

§ See e.g., 45 C.F.R. part 46, 21 C.F.R. part 56; 42 U.S.C. §241(d); 5 U.S.C. §552a; 44 U.S.C. §3501 et seq.

