## Supplemental materials and figures for "SARS-CoV-2 Seroprevalence Survey Among District Residents Presenting for Serologic Testing at Three Community-Based Test Sites — Washington, DC, July–August, 2020": Supplemental Materials File_DC Serosurvey Manuscript v10_2.15.21.pdf

**SUPPLEMENTARY MATERIALS FILE**

### METHODS

#### *Study Design and Population*

Sample size calculations were based on an estimated seroprevalence of 10%, a response rate of 50%, a design effect of 2.0, and a margin of error of 5%. Using a modified approach to the CDC Community Assessment for Public Health Emergency Response (CASPER) Geographic Information System (GIS) Toolbox, 56 census blocks in DC (based on 2010 U.S. Census data) were randomly selected without replacement and with probability proportional to number of occupied households [1]. The 30x7 sampling frame suggested in traditional CASPER methodology was modified to expand the number of randomly selected census blocks from 30 to 56 in order to use a higher margin of error while also increasing precision. Census blocks were selected using a random number generator, and those with more than 10 residential housing units were considered eligible for selection. One of the 56 randomly sampled census blocks had fewer than 10 residential household units and was subsequently re-sampled.

A multistep matching of the geocoded addresses of households within selected sample blocks was carried out. First, all potential residential premise addresses within the 56 randomly sampled census blocks were identified using property and address data from the Integrated Tax System Public Extract dataset and property use codes provided by the Government of the District of Columbia's Office of the Chief Financial Officer [2]. Residential property addresses were then geocoded using the Master Address Repository (MAR) Geocoder for DC. The MAR database was used to validate locations and identify apartment/condominium unit numbers. Household addresses were visually examined using Google Earth to ensure addresses represented residential properties. Fifteen households from each census block were randomly selected for participation using a random number generator. To account for incorrect and vacant addresses, as well as non-response, we expanded the original sampling frame from 10 to 15 households per census block to oversample by 50%. Consistent with the CDC CASPER toolkit, each household address had an equal chance of being selected.

A total of 839 households were initially sampled and contacted via mail. Reminder postcards were mailed to households that had not made a serology appointment by the second week of the study period. Response rates for the random sample were tracked in real time through daily monitoring of the serology appointment scheduling calendar, which was directly connected to the appointment call center. Due to the number of invitation letters returned to sender, an additional 40 household addresses were re-sampled and contacted via mail during week 3, yielding a total of 879 invited households. Forty-one (73%) of 56 randomly selected census blocks were represented, with an average of 1.8 households enrolled per census block (range: 0–7).

People who completed both the questionnaire and the blood draw were considered to be enrolled; people with invalid serologic test results (i.e., rejected specimens, lost samples, indeterminate results) or incomplete questionnaires were excluded from analyses.

#### *Participant Recruitment*

Invited residents were given a unique invitation code and were instructed to schedule an appointment through the appointment call center at one of three serology test sites operated by DC Health. The mailed invitation letter also included instructions for participating households to use their unique invitation code to obtain complimentary transportation to and from the testing sites through a contracted arrangement with DC Yellow Cab. Community advertising and outreach efforts included Mayor Bowser's Press Conferences, Mayor Bowser's Community Telephone Town Halls, media coverage and radio interviews, social media postings, Mayor Bowser's Coronavirus website, and community outreach with the Mayor's Office of Latino Affairs (MOLA) and sister agencies.

***Specimen Collection and Laboratory Analysis***

Three to four individually enclosed phlebotomy stations and one consolidated laboratory space were located within each site's mobile trailer. Each site's clinical team consisted of a head nurse, a medical technologist, and 3–4 phlebotomists. Serology test sites were open to invited participants and the general public on weekdays from 9am–3pm (Sites 1 and 2) and 10am–4pm (Site 3) during the first three weeks of enrollment. Two test sites remained open during week 4, with operating hours extended to 9am–7pm (Site 1) and 10am–8pm (Site 3).

DFS-PHL conducted an in-house validation of the LIAISON XL SARS-CoV-2 S1/S2 IgG assay by testing a panel of 36 sera samples (20 positive cases and 16 negative controls) commercially purchased from Plasma Service Group Inc. This in-house validation estimated the test sensitivity to be 90.0% (95% CI 77%, 100%) and estimated the test specificity to be 100% (95% CI 100%).

**References:**


1. Centers for Disease Control and Prevention (CDC), *Community Assessment for Public Health Emergency Response (CASPER) Toolkit: Third Edition*. 2019.

2. Office of the Chief Financial Officer (OCFO), *Integrated Tax System Public Extract*, in *Open Data DC*. 2020, Government of the District of Columbia Office of the Chief Financial Officer (OCFO): Washington, D.C. Available from: https://opendata.dc.gov/datasets/integrated-tax-system-public-extract/data.

Supplemental Figure 1: Timeline of SARS-CoV-2 Seroprevalence Survey in the District of Columbia

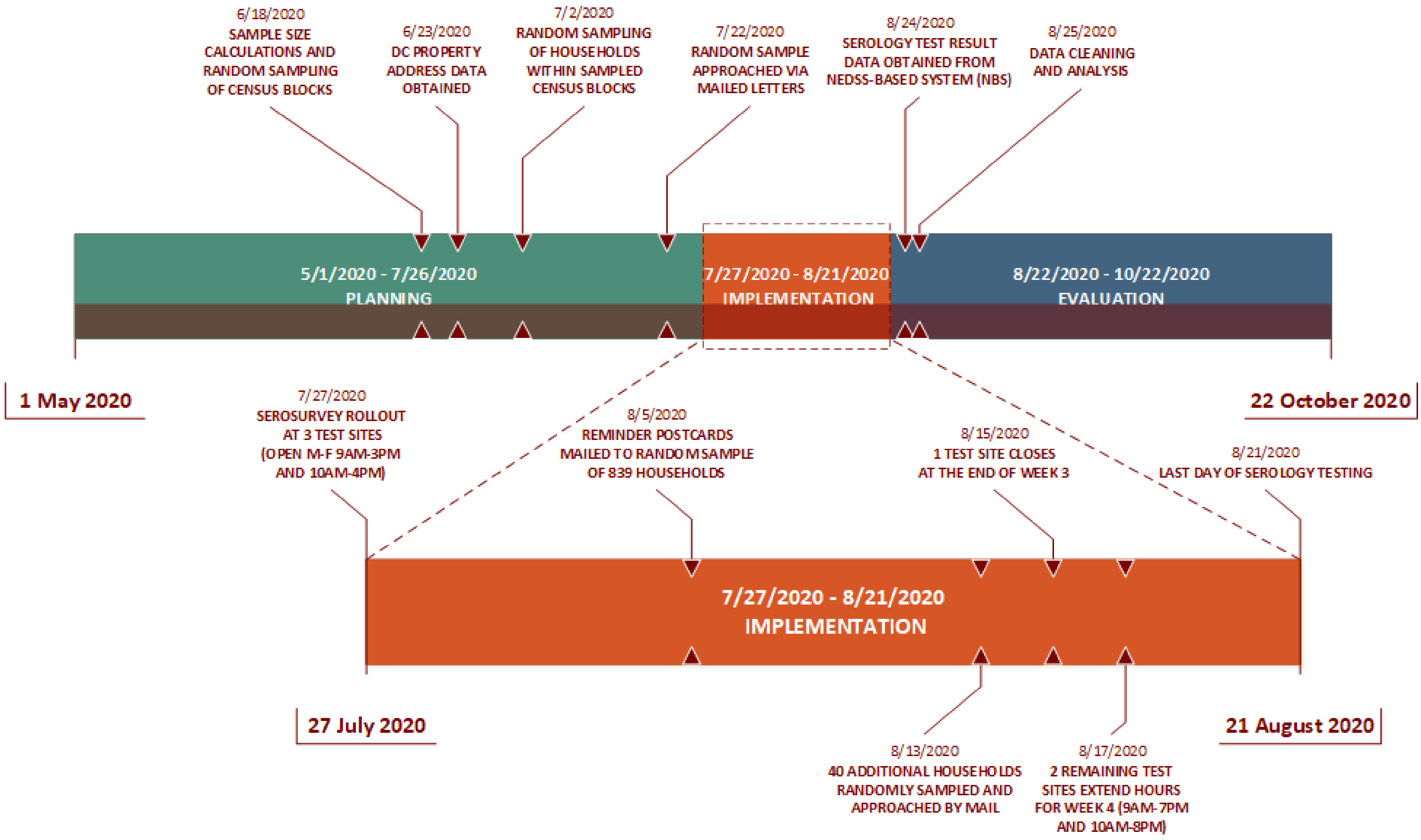

**Supplemental Figure 2: Geographic distribution of DC Health serology test sites and seroprevalence survey participants — Washington, DC, July 27–August 21, 2020**

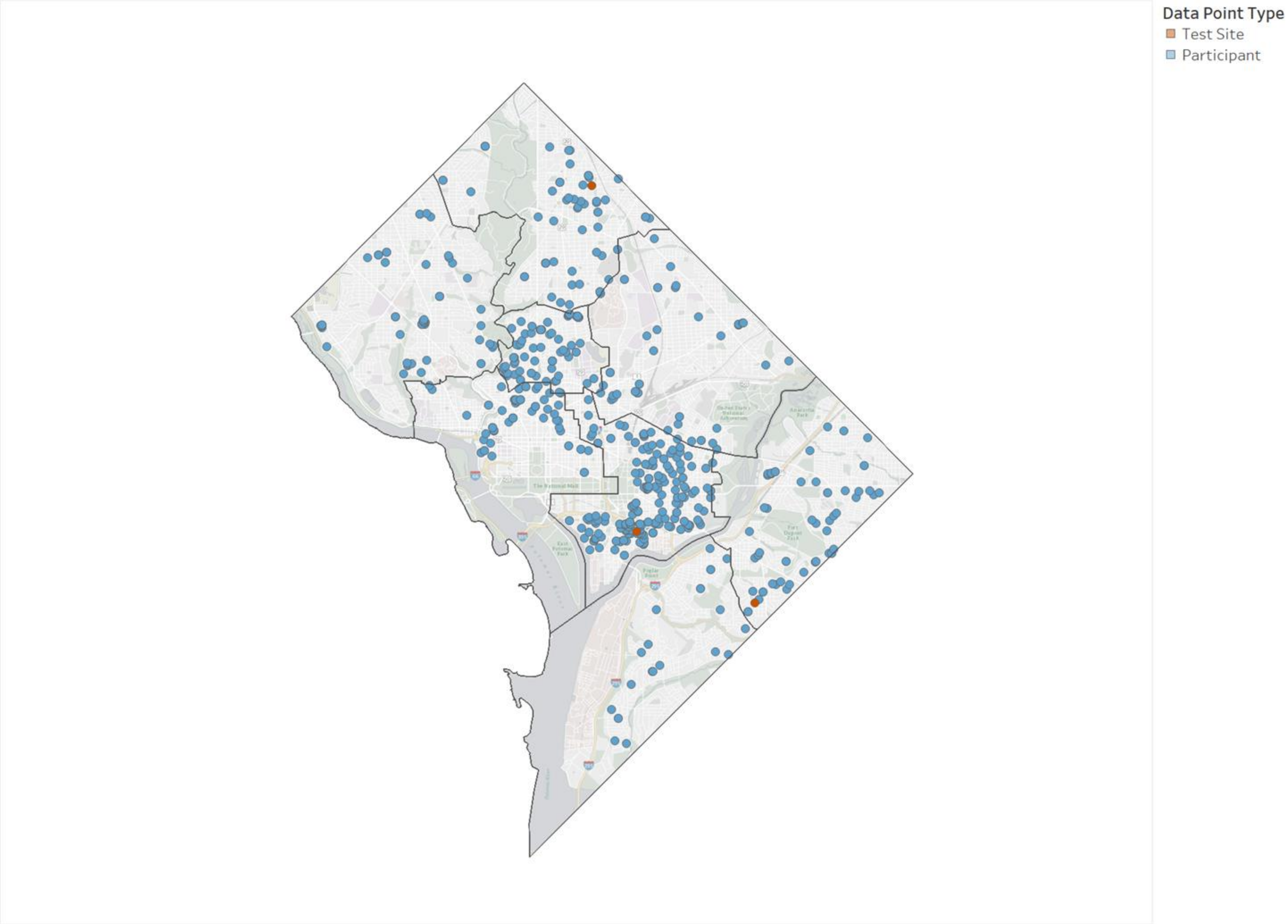


Map based on MAR Longitude and MAR Latitude and MAR Latitude. Color shows details about Data Point Type.

**Supplemental Figure 3: Geographic distribution of DC Health seroprevalence survey participants by ward of residence — Washington, DC, July 27–August 21, 2020**

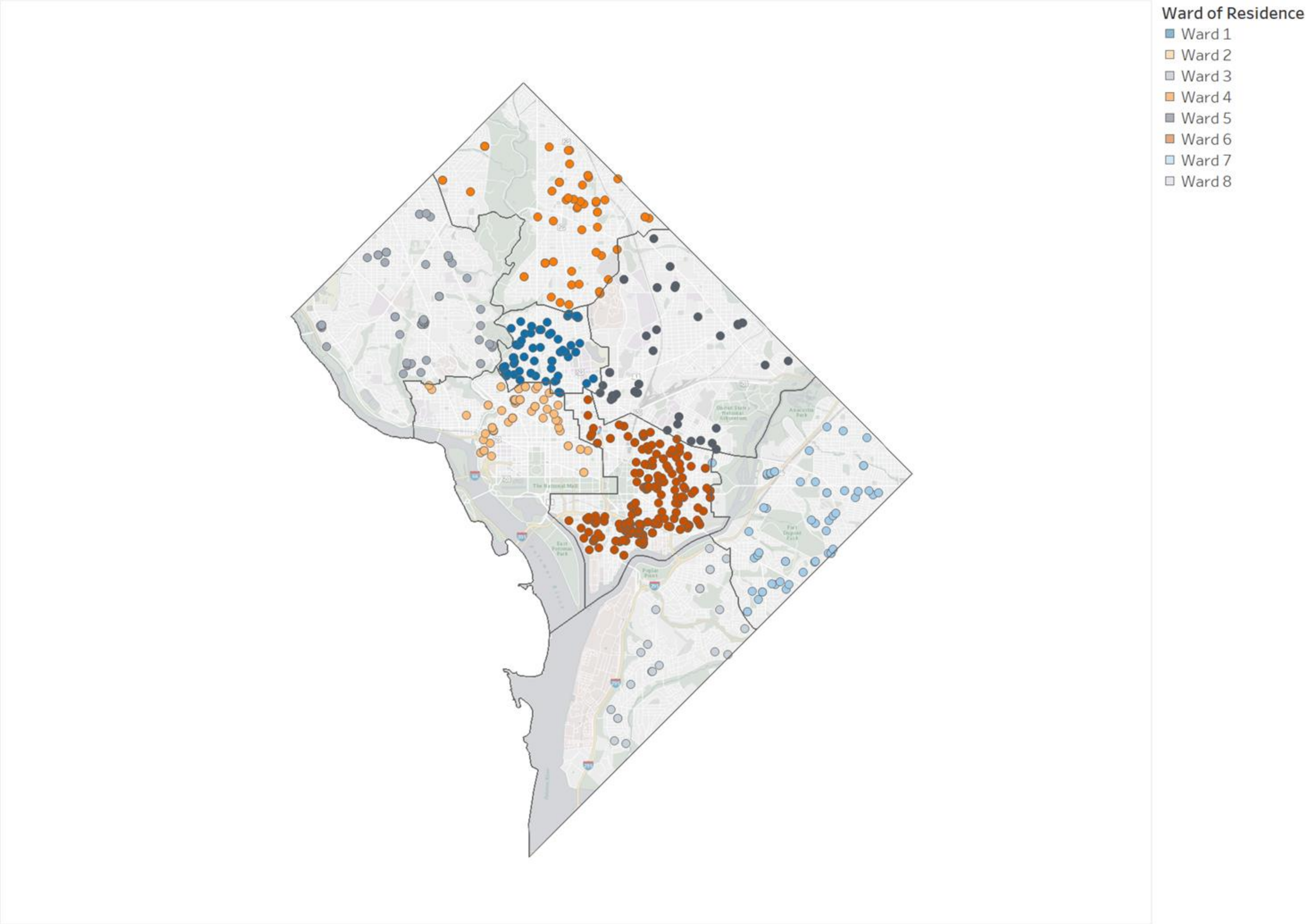


Map based on MAR Longitude and MAR Latitude and MAR Latitude. Color shows details about Ward of Residence. The data is filtered on Data Point Type, which keeps Participant.

**Supplemental Table 1: Characteristics of District residents tested for SARS-CoV-2 IgG antibodies by community-based seroprevalence survey stratified by sampling methodology — Washington, DC, July 27–August 21, 2020**

| Characteristic | Random Sample (N = 156) | | Convenience Sample (N = 515) | | p-value[¶] |
|---|---|---|---|---|---|
| | No. | Percent* (95% CI)[†] | No. | Percent* (95% CI)[†] | |
| ***Gender*** | | | | | |
| Female | 81 | 51.9 (43.8, 60.0) | 278 | 54.0 (49.6, 58.4) | 0.652 |
| Male | 75 | 48.1 (40.0, 56.2) | 237 | 46.0 (41.7, 50.4) | |
| ***Race/Ethnicity*** | | | | | |
| Hispanic ethnicity | 7 | 4.5 (1.8, 9.0) | 47 | 9.1 (6.8, 12.0) | 0.147 |
| Black, non-Hispanic | 39 | 25.0 (18.4, 32.6) | 102 | 19.8 (16. 5, 23.5) | |
| White, non-Hispanic | 98 | 62.8 (54.7, 70.4) | 305 | 59.2 (54.8, 63.5) | |
| Other, non-Hispanic | 9 | 5.8 (2. 7, 10. 7) | 43 | 8.4 (6.1, 11.1) | |
| Other, unknown | 3 | 1.9 (0.4, 5.5) | 18 | 3.5 (2.1, 5.5) | |
| ***Age Group (yrs)***[§] | | | | | |
| 6–17 | 4 | 2.6 (0.7, 6.4) | 25 | 4.9 (3.2, 7.1) | 0.016 |
| 18–29 | 28 | 18.0 (12.3, 24. 9) | 97 | 18.8 (15.6, 22.5) | |
| 30–49 | 67 | 43.0 (35.1, 51.1) | 254 | 49.3 (44.9, 53.7) | |
| 50–64 | 31 | 19.9 (13.9, 27.0) | 101 | 19.6 (16.3, 23.3) | |
| ≥65 | 26 | 16.7 (11.2, 23.5) | 38 | 7.4 (5.3, 10.0) | |
| ***Prior Testing for SARS-CoV-2*** | | | | | |
| Yes | 45 | 28.9 (21.9, 36.6) | 220 | 42.7 (38.4, 47.1) | 0.002 |
| ***COVID-Like Illness[§§] since January 1, 2020*** | | | | | |
| Yes | 31 | 19.9 (13.9, 27.0) | 173 | 33.6 (29.5, 37.9) | 0.001 |
| ***Medical History*** | | | | | |
| Any chronic condition[††] | 43 | 27.6 (20.7, 35.3) | 137 | 26.6 (22.8, 30.6) | 0.812 |
| Chronic lung disease | 27 | 17.3 (11.7, 24.2) | 74 | 14.4 (11.5, 17.7) | 0.369 |
| Cardiovascular disease | 16 | 10.3 (6.0, 16.1) | 65 | 12.6 (9.9, 15.8) | 0.427 |
| Chronic kidney disease | 3 | 1.9 (0.4, 5.5) | 5 | 1.0 (0.3, 2.3) | 0.397 |
| Liver Disease | 1 | 0.6 (0.0, 3.5) | 7 | 1.4 (0.6, 2.8) | 0.689 |

| | | | | | |
|---|---|---|---|---|---|
| Diabetes mellitus | 3 | 1.9 (0.4, 5.5) | 13 | 2.5 (1.4, 4.3) | 0.999 |
| Autoimmune, Rheumatologic, or Immunocompromising condition | 6 | 3.9 (1.4, 8.2) | 17 | 3.3 (1.9, 5.2) | 0.743 |
| Seasonal allergies | 83 | 53.2 (45.1, 61.2) | 285 | 55.3 (50.9, 59.7) | 0.639 |
| Recent/current pregnancy | 1 | 0.6 (0.0, 3.5) | 5 | 1.0 (0.3, 2.3) | 0.893 |
| ***Current Employment Status and Location*** | | | | | |
| Unemployed/furloughed | 29 | 18.6 (12.8, 25.6) | 66 | 12.8 (10.1, 16.0) | 0.005 |
| Employed outside the home | 25 | 16.0 (10.7, 22.7) | 121 | 23.5 (19.9, 27.4) | |
| Employed and teleworking | 69 | 44.2 (36.3, 52.4) | 265 | 51.5 (47.1, 55.9) | |
| Retired | 21 | 13.5 (8.5, 19.8) | 34 | 6.6 (4.6, 9.1) | |
| Student or <18 years of age | 12 | 7.7 (4.0, 13.1) | 29 | 5.6 (3.8, 8.0) | |
| ***Exposures since January 1, 2020*** | | | | | |
| No known exposures | 99 | 63.5 (55.4, 71.0) | 316 | 61.4 (57.0, 65.6) | 0.636 |
| Contact with ≥1 person with confirmed COVID-19 | 10 | 6.4 (3.1, 11.5) | 70 | 13.6 (10.8, 16.9) | 0.015 |
| Contact with ≥1 person with respiratory symptoms (not confirmed COVID-19) | 16 | 10.3 (6.0, 16.1) | 90 | 17.5 (14.3, 21.0) | 0.030 |
| International travel (outside the United States) | 37 | 23.7 (17.3, 31.2) | 79 | 15.3 (12.3, 18.8) | 0.015 |
| ***Number of Participating Household Members*** | | | | | |
| 1 | 59 | 37.8 (30.2, 45.9) | 322 | 62.5 (58.2, 66.7) | <0.0001 |
| 2 | 60 | 38.5 (30.8, 46.6) | 148 | 28.7 (24.9, 32.9) | |
| 3 | 12 | 7.7 (4.0, 13.1) | 27 | 5.2 (3.5, 7.5) | |
| 4 | 20 | 12.8 (8.0, 19.1) | 8 | 1.6 (0.7, 3.0) | |
| 5 | 5 | 3.2 (1.1, 7.3) | 10 | 1.9 (0.9, 3.5) | |

**Abbreviations:** IgG = Immunoglobulin G; DC = District of Columbia; No. = Number; CI = Confidence Interval;
*Percentages for mutually exclusive variables might not total 100.0 due to rounding.
[†]Represents exact 95% confidence intervals for the binomial proportions.

[§]Minimum age for serologic testing was 6 years old.
[¶]For categorical variables, Pearson's chi-square tests were performed where cell counts were sufficient and Fisher's exact test were carried out where any expected cell counts were less than 5.
[§§]An illness was categorized as compatible with COVID-19 if reported symptoms met the Council of State and Territorial Epidemiologists (CSTE) clinical criteria in the case definition, including (1) cough, difficulty breathing or shortness of breath, new loss of taste, or new loss of smell, or (2) two or more other symptoms (fever [measured or subjective], chills, rigors, myalgia, headache, sore throat, nausea or vomiting, diarrhea, fatigue, congestion or runny nose). https://cdn.ymaws.com/www.cste.org/resource/resmgr/ps/positionstatement2020/Interim-20-ID-02_COVID-19.pdf
[¶¶]Some participants reported multiple chronic conditions; chronic conditions included chronic lung diseases, cardiovascular diseases, chronic kidney diseases, liver diseases, diabetes mellitus, and autoimmune, rheumatologic, or immunocompromised conditions.

**Supplemental Table 2: Estimated seroprevalence of SARS-CoV-2 IgG antibodies among District residents enrolled in a community-based seroprevalence survey — Washington, DC, July 27–August 21, 2020**

| Group | Sampling Method | Sample Size | Available Data | Estimated Seroprevalence (95% CI)[†] |
|---|---|---|---|---|
| **Group 1** | Random Sample | 156 | Questionnaire and serology lab result | 3.2 (1.1, 7.3) |
| **Group 2** | Convenience Sample | 515 | Questionnaire and serology lab result | 8.9 (6.6, 11.7) |
| ***TOTAL*** | *Random and Convenience Samples* | *671* | *Questionnaire and serology lab result* | *7.6 (5.7, 9.9)* |

**Abbreviations:** IgG = Immunoglobulin G; DC = District of Columbia; CI = Confidence Interval;
[†]Represents exact 95% confidence intervals for the binomial proportions.
[§]Represents the final analytic sample (Groups 1 and 2 combined).

**Supplemental Table 3: SARS-CoV-2 Seroprevalence Questionnaire — Washington, DC, July 27–August 21, 2020**

| Variable / Field Name | Field Type | Field Label | Choices, Calculations, OR Slider Labels |
|---|---|---|---|
| ***BASIC INFORMATION*** | | | |
| surv_date | text | Record Creation Date | *Automatically generated* |
| person_completing | radio | Relation to individual completing this questionnaire: | 1, Self \| 2, Parent \| 3, Guardian \| 4, Caretaker |
| fname | text | First name of person completing the questionnaire: | |
| lname | text | Last name of person completing the questionnaire: | |
| fname_proxy | text | First name of household member you are completing this questionnaire on behalf of: | |
| lname_proxy | text | Last name of household member you are completing this questionnaire on behalf of: | |
| st_address | text | Household Street Address | |
| apt_address | text | Apartment/Unit/Suite | |
| zip | text | ZIP Code | |
| participantid | text | Participant ID: | |
| dob | text | Date of Birth: | |
| birthsex | radio | Sex Assigned at Birth: | 1, Male \| 2, Female |

| | | | |
|---|---|---|---|
| currentgender | radio | Current Gender Identity: | 1, Male \| 2, Female \| 3, Transgender M-F \| 4, Transgender F-M \| 5, Other gender not specified above \| 6, Prefer not to answer |
| ethnicity | radio | Ethnicity: | 1, Hispanic \| 2, Non-Hispanic \| 3, Unknown/Other |
| race | radio | Race: | 1, Black \| 2, White \| 3, Asian \| 4, Native Hawaiian or other Pacific Islander \| 5, American Indian or Alaska Native \| 6, Multi-racial \| 7, Unknown/Other |
| priortest | radio | Have you ever been tested for SARS-CoV-2 (the virus that causes COVID-19)? | 1, Yes \| 2, No \| 3, Don't know or can't remember |
| covtest_ct | text | How many times have you been tested for SARS-CoV-2 (the virus that causes COVID-19)? | |
| cs_or_rs | checkbox | How did you hear about this community-based antibody survey? | 1, Social Media \| 2, Mailed Letter \| 3, News \| 4, Friends/Family \| 5, Other |
| other_ad | notes | If other, please specify exactly how you came to learn of this community-based serology survey: | |
| survey_assent | radio | Do you agree to participate? | 1, YES - agree to complete the questionnaire and the blood draw \| 2, YES - agree to complete the questionnaire only \| 3, NO - do not agree to participate |
| decline_reason | radio | Please provide the reason why you do not wish to participate. | 1, Already had an antibody (blood) test done \| 2, Already had a diagnostic (throat/nasal/saliva) test done \| 3, Not interested \| 4, Too inconvenient \| 5, Other |
| decline_desc | notes | Please specify why you are declining to participate: | |
| ***INFORMATION ABOUT RECENT ILLNESS: You are now going to be asked a series of questions about your health status since January 1st of this year. If you were sick multiple times since January 1st, 2020, please answer the following series of questions based on the most recent time you were sick.*** | | | |
| illep1 | radio | Since January 1, 2020, have you experienced any of the following symptoms (anything different from any pre-existing conditions) for more than one day: cough, fever, chills, shortness of breath or difficulty breathing, fatigue, muscle or body aches, headache, new loss of taste or smell, sore throat, runny or stuffy nose, nausea, vomiting, or diarrhea? | 1, Yes \| 2, No \| 3, Don't know or can't remember |

| | | | |
|---|---|---|---|
| illep1_sxsst_date | text | When was the first day that you began to feel sick, or the first day you experienced symptoms? (Please refer to a calendar or smart phone to help recall the date) | |
| illep1_allsxs | checkbox | Which of the following symptoms have you experienced during this illness? Please check all that apply: | 1, Cough \| 2, Chills \| 3, Fever measured with thermometer \| 4, Felt warm/feverish \| 5, Runny/stuffy nose \| 6, Sore throat \| 7, Chest pain \| 8, Muscle pain \| 9, Difficulty breathing \| 10, Abdominal pain \| 11, Nausea \| 12, Vomiting \| 13, Diarrhea \| 14, Headache \| 15, Fatigue \| 16, New loss of taste \| 17, New loss of smell \| 18, Other |
| illep1_othersxs | text | If you have experienced other symptoms not listed above, please specify: | |
| illep1_maxtemp | text | What was your highest recorded temperature during this illness? Please provide the maximum temperature measured in degrees Fahrenheit (F) | |
| illep1_sxsend | radio | Have any of your symptoms improved since you began feeling sick? | 1, Yes \| 2, No \| 3, Don't know |
| illep1_duration | text | Counting that first day, how many total days were you sick until you started to feel much better and close to back to your usual health? (Please refer to a calendar or smart phone to help recall the duration) | |
| illep1_seekcare | radio | Did you go to a doctor, clinic, or emergency room because of this illness? | 1, Yes \| 2, No \| 3, Don't know or can't remember |
| illep1_caresetting | checkbox | Which type of care setting(s) did you use for this illness? (Please check all that apply) | 1, Clinic \| 2, Urgent Care \| 3, Hospital Emergency Room \| 4, Telemedicine Visit \| 5, Don't know or can't remember \| 6, Other |
| illep1_hosp | radio | Did you stay overnight in the hospital for this illness? | 1, Yes \| 2, No \| 3, Don't know or can't remember |
| illep1_hospdate | text | On which date were you admitted to the hospital for this illness? (Please refer to a calendar or smart phone to help recall the date) | |
| illep1_hospdays | text | How many days were you hospitalized for this illness? (If the exact number of days cannot be recalled, please provide an estimate) | |

| | | | |
|---|---|---|---|
| illep1_hosphcf | dropdown | Where were you hospitalized for this illness? | 1, Children's National Medical Center \| 2, George Washington University Hospital \| 3, Howard University Hospital \| 4, MedStar Georgetown University Hospital \| 5, MedStar Washington Hospital Center \| 6, MedStar National Rehabilitation Hospital \| 7, Sibley Memorial Hospital (John's Hopkins DC facility) \| 8, St. Elizabeth's Hospital \| 9, United Medical Center \| 10, Veteran's Affairs Medical Center \| 11, Other Healthcare Facility \| 12, Unknown |
| illep1_hosp_otherhcf | text | Please specify the name of the healthcare facility where you were hospitalized for this illness: | |
| illep1_anydiag | radio | Did you receive a diagnosis for this illness? | 1, Yes \| 2, No \| 3, Don't know or can't remember |
| illep1_diag | text | Please specify the diagnosis you received for this illness: | |
| illep1_covtest1 | radio | Were you tested for SARS-CoV-2 (the virus that causes COVID-19) for this illness? | 1, Yes - throat/nasal/saliva test \| 2, Yes - blood test \| 3, No \| 4, Don't know |
| illep1_covtest1_site | radio | Which body site was swabbed for COVID-19 testing? | 1, Nose \| 2, Throat \| 3, Saliva \| 4, Don't know or can't remember |
| illep1_covtest1_date | text | On which date were you tested for SARS-CoV-2 (the virus that causes COVID-19)? (Please refer to a calendar or smart phone to help recall the date) | |
| illep1_covtest1_result | radio | What was your result for this SARS-CoV-2 (COVID-19) test? | 1, Negative \| 2, Positive \| 3, Have not received test result \| 4, Don't know |
| illep1_covtest1rec | radio | Did a doctor or nurse evaluating you, or staff from the Health Department, recommend you get tested with a swab, or did you decide to get tested on your own? | 1, Recommended by doctor, nurse, or Health Department \| 2, Decided on my own \| 3, Don't know |
| illep1_covtest1reason | text | Why did you get tested? | |
| illep1_covtest1sxs | radio | Were you sick or having some symptoms around the time your swab was taken? | 1, Yes \| 2, No \| 3, Don't know or can't remember |
| illep1_covtest2 | radio | Were you tested for SARS-CoV-2 (the virus that causes COVID-19) a second time for this illness? | 1, Yes - throat/nasal/saliva test \| 2, Yes - blood test \| 3, No \| 4, Don't know |
| illep1_covtest2_site | radio | Which body site was swabbed for COVID-19 testing? | 1, Nose \| 2, Throat \| 3, Saliva \| 4, Don't know or can't remember |

| | | | |
|---|---|---|---|
| illep1_covtest2_date | text | On which date were you tested for SARS-CoV-2 (the virus that causes COVID-19) a second time? (Please refer to a calendar or smart phone to help recall the date) | |
| illep1_covtest2_result | radio | What was your result for this SARS-CoV-2 (COVID-19) test? | 1, Negative \| 2, Positive \| 3, Have not received test result \| 4, Don't know |
| illep1_covtest2rec | radio | Did a doctor or nurse evaluating you, or staff from the Health Department, recommend you get tested with a swab, or did you decide to get tested on your own? | 1, Recommended by doctor, nurse, or Health Department \| 2, Decided on my own \| 3, Don't know |
| illep1_covtest2reason | text | Why did you get tested? | |
| illep1_covtest2sxs | radio | Were you sick or having some symptoms around the time your swab was taken? | 1, Yes \| 2, No \| 3, Don't know or can't remember |
| illep1_covtest3 | radio | Were you tested for SARS-CoV-2 (the virus that causes COVID-19) a third time for this illness? | 1, Yes - throat/nasal/saliva test \| 2, Yes - blood test \| 3, No \| 4, Don't know |
| illep1_covtest3_site | radio | Which body site was swabbed for COVID-19 testing? | 1, Nose \| 2, Throat \| 3, Saliva \| 4, Don't know or can't remember |
| illep1_covtest3_date | text | On which date were you tested for SARS-CoV-2 (the virus that causes COVID-19) a third time? (Please refer to a calendar or smart phone to help recall the date) | |
| illep1_covtest3_result | radio | What was your result for this SARS-CoV-2 (COVID-19) test? | 1, Negative \| 2, Positive \| 3, Have not received test result \| 4, Don't know |
| illep1_covtest3rec | radio | Did a doctor or nurse evaluating you, or staff from the Health Department, recommend you get tested with a swab, or did you decide to get tested on your own? | 1, Recommended by doctor, nurse, or Health Department \| 2, Decided on my own \| 3, Don't know |
| illep1_covtest3reason | text | Why did you get tested? | |
| illep1_covtest3sxs | radio | Were you sick or having some symptoms around the time your swab was taken? | 1, Yes \| 2, No \| 3, Don't know or can't remember |
| illep1_covtest4 | radio | Were you tested for SARS-CoV-2 (the virus that causes COVID-19) a fourth time for this illness? | 1, Yes - throat/nasal/saliva test \| 2, Yes - blood test \| 3, No \| 4, Don't know |
| illep1_covtest4_site | radio | Which body site was swabbed for COVID-19 testing? | 1, Nose \| 2, Throat \| 3, Saliva \| 4, Don't know or can't remember |
| illep1_covtest4_date | text | On which date were you tested for SARS-CoV-2 (the virus that causes COVID-19) a fourth time? (Please | |

| | | | |
|---|---|---|---|
| | | refer to a calendar or smart phone to help recall the date) | |
| illep1_covtest4_result | radio | What was your result for this SARS-CoV-2 (COVID-19) test? | 1, Negative \| 2, Positive \| 3, Have not received test result \| 4, Don't know |
| illep1_covtest4rec | radio | Did a doctor or nurse evaluating you, or staff from the Health Department, recommend you get tested with a swab, or did you decide to get tested on your own? | 1, Recommended by doctor, nurse, or Health Department \| 2, Decided on my own \| 3, Don't know |
| illep1_covtest4reason | text | Why did you get tested? | |
| illep1_covtest4sxs | radio | Were you sick or having some symptoms around the time your swab was taken? | 1, Yes \| 2, No \| 3, Don't know or can't remember |
| illep1_absences | radio | Did you miss any days of school or work because of this illness? | 1, Yes \| 2, No \| 3, Don't know or can't remember |
| illep1_abscencect | text | How many days of work or school did you miss because of this illness? | |
| illep1_publicfreq | radio | How often did you go to public places (e.g., school, work, store, place of worship) during the 7 days after this illness began? (Please do not include visits to the doctor) | 1, Multiple times per day \| 2, Once per day \| 3, Several times during the 7-day period \| 4, Rarely \| 5, Never \| 6, Don't know or can't remember |
| illep1_visitors | radio | Did any family or friends come over to visit you during this illness? | 1, Yes \| 2, No \| 3, Don't know or can't remember |
| ***INFORMATION ABOUT MEDICAL HISTORY: You are now going to be asked a series of questions about your past and current medical history.*** | | | |
| pmhx_allergies | radio | Do you have seasonal allergies? | 1, Yes \| 2, No \| 3, Don't know |
| pmhx_lung | checkbox | Do you have any of the following chronic lung diseases? Please check all that apply: | 1, Asthma/reactive airway disease \| 2, Chronic Obstructive Pulmonary Disease (COPD) \| 3, Emphysema \| 4, Other \| 5, None \| 6, Don't know |
| pmhx_lungoth | text | Please specify the other chronic lung disease(s): | |
| pmhx_diabetes | radio | Do you have Diabetes Mellitus? | 1, Yes \| 2, No \| 3, Don't know |
| pmhx_heart | checkbox | Do you have any of the following cardiovascular (heart) diseases? Please check all that apply: | 1, Hypertension \| 2, Stroke \| 3, Coronary artery disease \| 4, Heart failure/Congestive heart failure \| 5, Congenital heart disease \| 6, Other \| 7, None \| 8, Don't know |
| pmhx_heartoth | text | Please specify the other cardiovascular (heart) diseases: | |
| pmhx_kidney | checkbox | Do you have any of the following renal (kidney) diseases? Please check all that apply: | 1, Chronic kidney disease \| 2, Renal failure/dialysis \| 3, Other \| 4, None \| 5, Don't know |
| pmhx_kidneyoth | text | Please specify the other renal (kidney) diseases: | |

| | | | |
|---|---|---|---|
| pmhx_liver | checkbox | Do you have any of the following liver diseases? Please check all that apply: | 1, Hepatitis A \| 2, Hepatitis B \| 3, Hepatitis C \| 4, Cirrhosis \| 5, Other \| 6, None \| 7, Don't know |
| pmhx_liveroth | text | Please specify the other liver diseases: | |
| pmhx_immunocomp | radio | Are you immunocompromised? | 1, Yes \| 2, No \| 3, Don't know |
| pmhx_immuno | checkbox | Which of the following conditions do you have? | 1, HIV infection \| 2, AIDS or CD4 count < 200 \| 3, Solid organ transplant \| 4, Stem cell transplant \| 5, Lupus \| 6, Rheumatoid Arthritis \| 7, Multiple sclerosis \| 8, Cancer (current/in treatment or diagnosed in last 12 months) \| 9, Other \| 10, None \| 11, Don't know |
| pmhx_immunoth | text | Please specify the other condition: | |
| recentpreg | radio | Are you currently pregnant or have you had a child within the last 6 weeks? | 1, Yes \| 2, No \| 3, Don't know \| 4, Not applicable |
| ***INFORMATION ABOUT POTENTIAL EXPOSURES: You are now going to be asked a series of questions about your potential exposures to SARS-CoV-2 (the virus that causes COVID-19).*** | | | |
| employ | radio | Please describe your current employment status: | 1, Employed - currently working outside of the house some days or everyday \| 2, Employed - teleworking every day that I work \| 3, Employed - furloughed or lost job since outbreak started \| 4, Not applicable - full-time student or under 18 years old \| 5, Not employed \| 6, Retired |
| essential | radio | Do you work at a place that is considered an "essential service"? | 1, Yes \| 2, No \| 3, Don't know |
| essential_job | radio | What type of essential service field do you work in: | 1, Healthcare facility \| 2, Grocery store \| 3, Restaurant \| 4, Non-grocery store \| 5, Home health-aid/care-giver \| 6, Pharmacy \| 7, Warehouse/shipping center \| 8, Government/public service \| 9, Delivery driver, parcel (e.g., USPS, UPS, FedEx) \| 10, Delivery driver, food (e.g., grocery, restaurant, Uber Eats) \| 11, Public transportation/airline/airport \| 12, Other |
| essential_oth | text | Please specify the other type of essential service field that you work in: | |
| contact | radio | Have you had contact with anyone diagnosed with a confirmed SARS-CoV-2 infection (also called COVID-19) while they were sick? | 1, Yes - 1 person \| 2, Yes - more than one person \| 3, No \| 4, Don't know |

| | | | |
|---|---|---|---|
| caserelation | checkbox | What is your relationship to the person(s) with a confirmed SARS-CoV-2 infection (COVID-19)? Please check all that apply: | 1, Spouse/Partner \| 2, Child \| 3, Parent \| 4, Other household relatives \| 5, Friend \| 6, Healthcare worker \| 7, Co-worker \| 8, Classmate \| 9, Roommate \| 10, Patient \| 11, Client \| 12, Contact only - no relationship \| 13, Other |
| caserelation_oth | text | Please specify your relationship(s) to the sick person(s): | |
| illcontact | radio | Have you had contact with anyone who was sick with respiratory symptoms but was NOT diagnosed with a confirmed SARS-CoV-2 infection (COVID-19) while they were sick? | 1, Yes \| 2, No \| 3, Don't know |
| illrelation | checkbox | What is your relationship to the sick person(s)? Please check all that apply: | 1, Spouse/Partner \| 2, Child \| 3, Parent \| 4, Other household relatives \| 5, Friend \| 6, Healthcare worker \| 7, Co-worker \| 8, Classmate \| 9, Roommate \| 10, Patient \| 11, Client \| 12, Contact only - no relationship \| 13, Other |
| sickrelation_oth | text | Please specify your relationship(s) to the sick person(s): | |
| anytravel | radio | Have you traveled out of the country since January 1, 2020? | 1, Yes \| 2, No \| 3, Don't know or can't remember |
| travel_desc | notes | Please provide the specific countries you have visited since January 1, 2020, and the corresponding dates of travel: | |
| ***NEXT HOUSEHOLD MEMBER*** | | | |
| add_individual | yesno | Has an additional individual within your household agreed to participate in the survey? | 1, Yes \| 2, No |
