## Supplementary figures and images for "SARS-CoV-2 Seroprevalence Survey Among District Residents Presenting for Serologic Testing at Three Community-Based Test Sites — Washington, DC, July–August, 2020"

### Figure 1 Manuscript v10_2.14.21.pdf

**Figure 1: DC Health Community Seroprevalence Survey Participant Flow Chart**

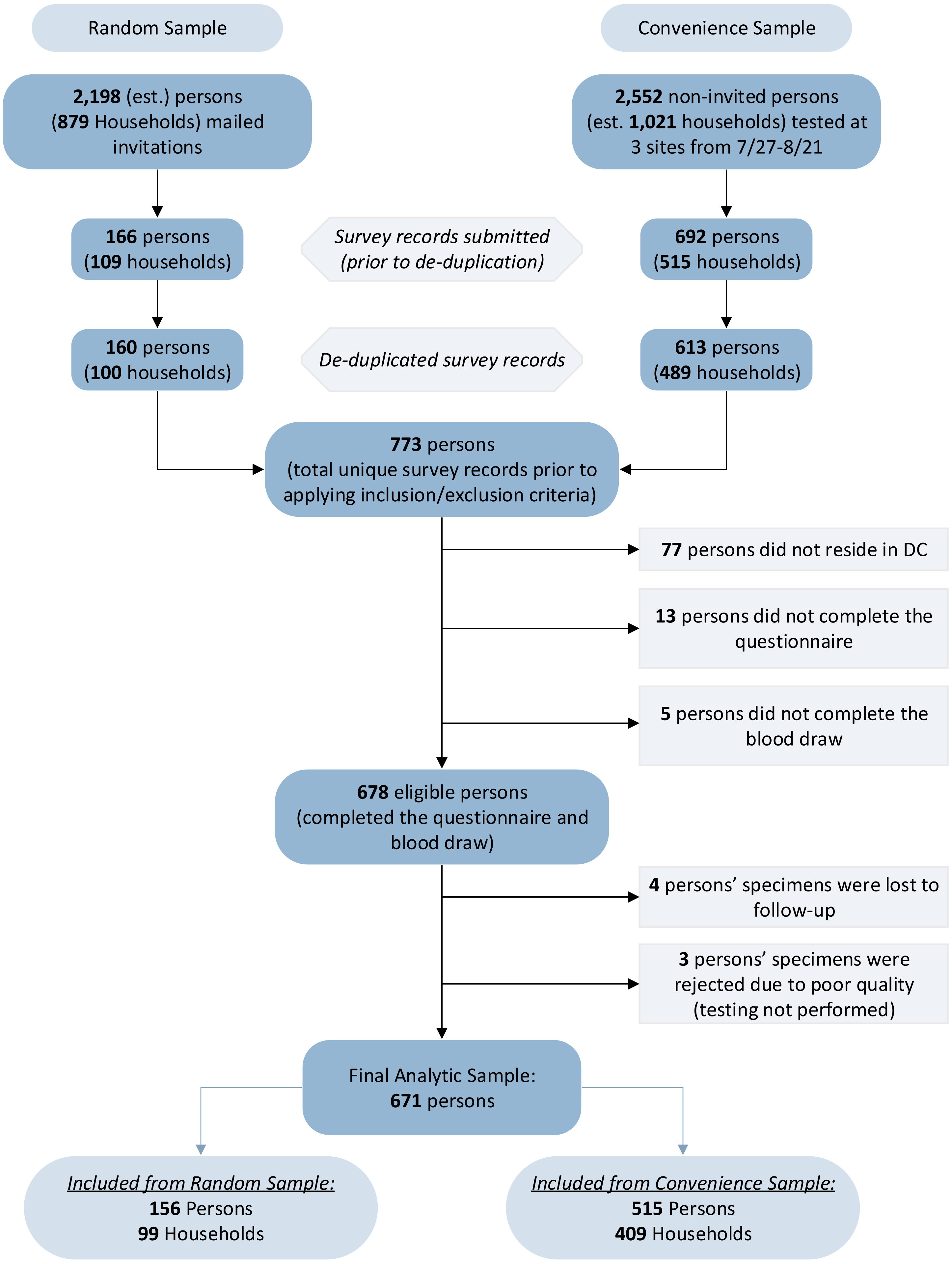
